# Aerodigestoscopy (ADS): A retrospective examination of the feasibility, safety, and comfort of a new procedure for the evaluation of physiological disorders of the aerodigestive tract

**DOI:** 10.1101/2022.08.16.22278864

**Authors:** Robert J. Arnold, Nina Bausek, Christopher S. Gaskill, Lawrence F. Johnson, Sigfredo Aldarondo, Cody Aull, Malik Midani, Tarek Midani, Ramiz Midani, Ashton S. Brown, Alana Wallace

## Abstract

Limited access to comprehensive assessment of physiological disorders of the upper aerodigestive tract by different specialties represents a barrier to care in rural healthcare settings, which may lead to misdiagnosis, underdiagnosis, and increased associated mortality. No single differential diagnostic exam for the comprehensive assessment of all functions of the upper aerodigestive tract is available to bridge this gap. We present a single procedure for assessment of physiological functions of the upper aerodigestive tract including swallow, voice, respiration, and cough while also screening for gastric retention and obstructive sleep apnea which might contribute to these disorders. Our procedure, called Aerodigestoscopy (ADS), represents a procedure incorporating other established exams as integral components to evaluate patients with aerodigestive disorders. Patients who might particularly benefit from our ADS diagnostic algorithm are those in a rural community referred by other health professionals due to complaints of aerodigestive symptoms and signs. We present a brief overview of how the procedure was developed, what it includes, and retrospective safety data from over 18,000 ADS procedures performed over the last 10 years, demonstrating safety and feasibility of this procedure.

## Introduction

Our experience in rural healthcare in the United States of America including community hospitals, physician offices, outpatient clinics, skilled nursing facilities, and home health agencies is that many of these rural healthcare settings do not have cost-effective and timely access to instrumental assessments for physiological disorders of the upper aerodigestive tract. To further confound the problem, simple symptoms do not imply simple disorders and a single professional’s knowledge base unfortunately does not cover all physiological disorders of the aerodigestive tract (AT). Although there are clinical disciplines which overlap, evidence suggests no single clinical discipline professes dominance for the diagnosis and treatment of all physiological disorders of the AT.

These various overlapping medical, surgical, and therapeutic disciplines inherently approach the process of diagnosing physiological disorders of the AT from their unique perspectives. Therefore, any single diagnostician is likely to focus solely on the chief complaint(s) most relevant to their discipline, risking a myopic view which may fail to identify significant comorbidities, and in turn may lead to an increased likelihood of misdiagnosis, missed diagnoses, costly inappropriate referrals to other professionals, increased morbidity, or even death while deferring the most appropriate treatment. Unfortunately, this reality exacerbates when patients with physiological disorders of the AT face limited access to aerodigestive professionals in rural settings. This persistent healthcare conundrum in rural settings resulted in the demand by rural physicians and speech-language pathologists (SLPs) to develop a single differential diagnostic instrumental assessment for the comprehensive examination of all physiological functions of the AT. This motivation forged our concept of the Aerodigestoscopy (ADS) procedure.

The ADS procedure is designed to simultaneously evaluate all aerodigestive dysfunctions in a single exam which include, but are not necessarily limited to swallow disorders (dysphagia) [1–4], motor speech disorders (dysarthria and apraxia of speech) [5–7], voice disorders [8–10], resonance disorders [11,12], cough disorders [13–15], breathing disorders [13,16–18], obstructive sleep disorders [19,20], and regurgitation disorders (dysemesis, backflow, and reflux) [21–23]. Impaired neuromuscular function of one part of the AT can result in a series of physiological dysfunctions elsewhere. For example, while impaired glottic closure can result in impaired voicing, it may also adversely affect the mechanical function of the thoracic cage muscles. The subsequent physiological deficits of the latter can result in further physiological impairments of defecation, micturition, and parturition which can be thought of as tertiary deficits. Physiological disorders of the upper aerodigestive tract can in turn lead to dehydration, malnourishment, weakness of the extremities, and generalized weakness with secondary issues of altered biochemistry, neurochemistry, cardiac chemistry, etc. Psychosocial and societal consequences of physiological disorders of the aerodigestive tract can include the compromise of mental health with depression and/or anxiety, and occupational consequences [24].

In both optometry and audiology, similar differential algorithm-driven diagnostic procedures with specific subcomponents are shared between physician and non-physician specialists as eye exams and hearing exams. Unfortunately, in the world of diagnosing physiological disorders of the AT, there is no shared unified algorithm between aerodigestive professionals. However, in medically complicated cases involving diagnosis and treatment of physiological disorders of the AT, a shared differential diagnostic algorithm between aerodigestive professions can save time and resources in addition to preserving the hope of a given patient who is waiting for a definitive diagnosis and treatment. In addition, such a shared diagnostic algorithm may optimize professional-to-professional communication.

The origin of ADS in 1989 resulted from the first author’s desire to create a procedure with robust diagnostic yield across the spectrum of physiological disorders of the aerodigestive tract. Its development began by surveying the various clinical and instrumental technologies and approved standards for the evaluation of various physiological disorders of the AT at that time. As the list of procedures began to grow, the next question posed was which of these procedures were capable of being integrated into a complementary unified differential diagnostic algorithm as sub-components with software guidance to evaluate each key physiological function of the upper aerodigestive tract. This task required software (ADS Software) to be developed in order to guide the holistic assessment in a time efficient manner while facilitating a seamless, intuitive movement through the administration of the procedure protocol. These activities included data collection, analysis, and evidence-based recommendations for further diagnostics, consultation by the most appropriate overlapping aerodigestive professional, and/or treatment(s).

While statistics pertaining to the collective incidence and prevalence of physiological disorders of the AT across the disciplines of therapy, nursing, medicine, and surgery are lacking, there are some reports giving insight to the incidence and prevalence of some disorders of the AT. Swallow disorders have a prevalence of 3% of the adult population, thus affecting more than 9 million people in the US, and causing over 60,000 deaths per year in the stroke population[25,26]. However, prevalence in the elderly may be significantly higher, with a reported average of 15%, and up to 60% in nursing homes [27,28]. The economic burden of dysphagia has been estimated to amount to $7 billion added to annual healthcare costs[25]. The observed mortality rate may be in part due to delayed or inaccurate diagnosis that overlooks significant comorbidities, resulting from the use of segmental instead of comprehensive diagnostic procedures [28,29]. Less information exists regarding the incidence and prevalence of voice disorders and motor speech disorders. Dysphonia has been shown to affect nearly one third of all persons across the lifespan [30]. Motor Speech Disorders (MSD) which include both dysarthria and apraxia of speech, a recent study reported by the Speech Pathology Section of the Mayo Clinic, MSDs account for about one-half of the primary communication disorder in people who present with neuropathology whereas at the same clinic MSDs represent the primary communication disorder in over one-third of all persons with an acquired disease of any type [31]. Gastroesophageal reflux disease (GERD) in North America has a reported prevalence of 18.1% to 27.8% [32]. The prevalence of obstructive sleep apnea (OSA) in the United States of America has been reported in the range of 3 to 7% [33].

The purpose of this paper is to examine the feasibility, safety, and comfort of the ADS procedure, present reasons for referral, and highlight the procedure’s diagnostic yields.

## Methods

**Category 1 Materials** = Durable Medical Equipment

- JedMed 58 cm Extended Length Nasopharyngoscope
- JedMed M-Camera
- JedMed Medical EDA audiovisual capture software
- ADS Software ver. 1.0 developed by Southeastern Biocommunication Associates, LLC
- Laptop computer with Windows operating system
- Sphygnomonometer
- Stethoscope
- Dual Pulse Oximeter-Capnographer
- Micro I Diagnostic Spirometer for Peak Expiratory Flow measurement.
- Micro Respiratory Pressure Meter for Maximum Inspiratory Pressure (MIP) & Maximum Expiratory Pressure (MEP).
- Calibrated Microphone

**Category 2 Materials** = Perishables & single use supplies.

- Infection control supplies
- Sample food and liquid consistencies, clothes protectors, napkins, towels, utensils, surgical lubricant, etc.

## Procedure

### P1 = Pre-ADS History & Physical

Set up of the ADS equipment and the food stimulus items are performed according to infection control protocol(s). The ADS software guides the clinician through the collection of some data prior to transnasal passage of the scope, so the clinician can be aware of the conditions of other aerodigestive functions before performing the endoscopic exam. First, the patient’s medical history is reviewed, including the reason for the referral for ADS, with particular emphasis on any diagnosis related to the AT. A physical exam is then conducted, including the following components: oromotor and facial exam (focusing on both morphology and cranial nerve function), neck exam and thoracic exam, otoscopy, initial vital signs (blood pressure, pulse, respiratory rate, and indirect oxygen saturation), and pulmonary function tests (PFTs) as indicated. Also, the current route for nutrition, hydration, and medication (e.g. oral intake, nasogastric tube, PEG tube, etc.) as well as any oral intake restrictions in terms of food, liquid, and medication consistencies are noted. Additionally, the STOP-BANG can be administered to screen for OSA [34]. In case of the presence of a head and/or neck tumor, the TNM staging system data is entered as appropriate when available [35].

### P2 = Perceptual Analysis of Voice

A perceptual analysis of vocal quality is performed, using the 4-point GRBAS scale (rating overall vocal severity, roughness, breathiness, asthenia, and strain)[36], while the ADS software records the audio sample. Additional specific perceptual descriptors (e.g. diplophonia, glottal fry, pItch breaks, tremor, etc) can also be reported here to enhance the perceptual analysis of voice.

### P3 = Acoustic Analysis of Voice

An acoustic analysis of voice is performed. Measurements include: fundamental frequency in Hz, vocal intensity in dB SPL (along with maximum phonation time (MPT) and cepstral peak prominence (CPP). All measurements are made with both a sustained vowel and a short connected speech sample. All measurements are taken in a quiet exam room while using a calibrated microphone placed at a fixed distance from the patient’s mouth using high-definition audio recording via the ADS software [37].

### P4 = Spectrographic Analysis of Voice

The spectrographic analysis of voice is plotted by the ADS software for visual analysis in the corresponding area of the ADS report for both voice samples taken during P3 [37].

### P5 = Initial Oral Exam

The endoscope is utilized to examine the oral cavity as it appears prior to the initiation of p.o. trials, and any abnormal findings are documented.

NOTE: A supplemental ADS flowchart as well as an anatomical depiction of the general position of the distal tip of the endoscope for each component of the ADS entitled “ADS Pathway” are found in the supplemental materials to enhance the description of the ADS procedure for P5 through P17.

### P6 = Anterior Nasal Exam

The endoscope is gently introduced to each naris to examine each side of the nasal cavity to determine any anatomical differences which may pose challenges for safe passage of the flexible fiberoptic endoscope. Additionally, relevant signs consistent with disorders, diseases, or physiological dysfunction are noted. The use of nasal saline, surgical lubricant, and oxymetazoline HCl, can be considered as needed.

### P7 = Nasopharyngeal Exam

As the scope is passed to the nasopharynx, imaging of the orifices of the Eustachian tubes as well as the adenoidal pad is performed, noting any abnormalities.

### P8 = Velopharyngeal Exam

The flexible fiberoptic endoscope is then utilized to complete an examination of velopharyngeal function. This subcomponent of the ADS procedure contains speech tasks to be performed to rule out velopharyngeal insufficiency (VPI), which can adversely impact nasal resonance, and swallow tasks to rule out velopharyngeal dysfunction (VPD). VPD can result in foods, liquids, and/or oral medications entering the posterior nasal cavity as well as result in a decrease of pharyngeal pressure and subsequent esophageal pressure for functional bolus passage. Signs of erythema, edema, and drainage from the adenoidal pad consistent with seasonal allergy(ies) are noted. In addition, any erythema, edema, or cobblestoning of the mucosa ascending from the pharynx into the inferior nasopharynx consistent with exposure to gastric acids are noted.

### P9 = Reflux Finding Score (RFS)

The endoscope is now advanced further into the pharynx to a position superior to the laryngeal vestibule and one or more images are taken to be analyzed by using the Reflux Finding Score (RFS) for the detection of signs characteristic of laryngopharyngeal reflux (LPR) [23].

### P10 = Stroboscopic Analysis of Voice

For the stroboscopic analysis of voice, the ADS procedure utilizes the Voice-Vibratory Assessment with Laryngeal Imaging (VALI) [38]. The VALI provides a framework for comprehensive stroboscopic examination of the larynx. It is ideal to perform the stroboscopic component of the ADS procedure prior to exposing the larynx and pharynx to food and liquid materials which may hinder full visualization of the glottis.

### P11 = Pharyngeal & Laryngeal Exam Under Constant Light prior to the initiation of p.o. trials

For this portion of the procedure, a gross assessment of the pharyngeal and laryngeal musculature is performed without p.o. trials. This can help to modify or develop new clinical hypotheses before the p.o. trials portion of the ADS procedure. This section begins with a physical screening for anatomical signs which may place an individual at risk for a pharyngeal swallow disorder, a voice disorder, OSA, etc.. In addition, it includes gross evaluation of the movement of some key structures of the larynx and pharynx (eg vocal fold adduction, abduction, pharyngeal contraction and shortening, etc).

### P12 = Transesophageal Passage and the Gastric Screen

Complete assessment of physiological functions of the upper aerodigestive tract requires an esophageal physiology exam as well as a gastric screening procedure. The gastric screen portion of this algorithm is based upon a four hour gastric emptying procedure utilized in nuclear medicine [39,40]. The gastric screen required the patient to be held *nil per os* (NPO) meaning nothing by mouth except for required oral medications, small volume medically necessary snack, or water, for at least 4 hours before the initiation of the ADS procedure. When possible, the Gastric Screen portion of the ADS is performed prior to the initiation of p.o. Trials. Prior to the passage of the endoscope through the cervical esophagus, a given patient’s non-oral versus oral status is considered. Once the endoscope is inside of the cervical esophagus, the clinician visualizes the appearance of the mucosa of the esophagus while advancing inferiorly to the lower esophageal sphincter (LES). The gastric screen portion is considered to be passed when the patient’s stomach is less than half full and failed when the patient’s stomach is found to be greater than half full. When a patient fails the gastric screen, then the clinician documents the finding of signs characteristic of abnormal gastric retention versus impaired gastric emptying. An abnormal physiological diagnosis of gastroesophageal dysphagia is made when the patient fails the gastric screen. An abdoöinal X-ray versus CT of the abdomen is recommended. Depending on the findings, a GI physician consultation is recommended.

### P13 = Esophageal Exam

NOTE: A table entitled “Esophageal Dysphagia: Helpful terms and concepts” as well as a table entitled “Regurgitation Disorders: Helpful terms and concepts” can be found in the supplemental material, both of which contain definitions to distinguish between these dysphysiological diagnostic terms as they relate to ADS esophageal findings.

#### LES Achalasia Screening

Once the gastric screen is completed, then the clinician can elect to start initial p.o. trials with the distal tip of the endoscope positioned in the pharynx versus positioned in the distal esophagus. Which portion of the aerodigestive tract to assess first with p.o. trials largely depends upon the data collected at step P1 when completing the Pre-ADS history and physical. Assuming the clinician elects to initiate p.o. trials in the esophagus, the endoscope is typically withdrawn to a level approximately 2 to 4 cm above the LES. Any anatomical irregularities of the LES and distal esophagus are described. It is desirable to screen for achalasia of the LES prior to the administration of any solid consistency foods. To screen for signs of achalasia of the LES, liquid trials may be initiated in 5 to 10 cc bolus sizes with the given patient’s current tolerated liquid consistency. In the event the total volume of the liquid consistency approaches 2 to 3 ounces while remaining mostly above the LES for longer than 5 minutes, then achalasia of the LES is suspected. This endoscopic screening methodology for detecting achalasia at the level of the LES is predicated upon an upright timed barium esophagram procedure [41,42]. In this case, the clinician makes a dysphysiological diagnosis of neurogenic gastroesophageal dysphagia characterized by signs consistent with achalasia of the LES. Referral is made to gastroenterology for appropriate tests to definitively rule in or rule out it’s presence (e.g. EGD with retroversion while inside the stomach to definitively rule out the presence of a mass in the fundus, high resolution manometry to definitively rule out failure of the LES to relax, etc.).

#### Motility Screening

After completing the screen for signs consistent with achalasia of the LES, the clinician notes the presence of any significant delay in the esophageal emptying of the liquid characteristic of primary and/or secondary dysmotility of the esophagus [43,44]. Any tertiary contractions are also noted. When the gastric screen and the LES achalasia screen are passed, any esophageal emptying differences related to impaired peristalsis would warrant a dysphysiological diagnosis of neurogenic esophageal dysphagia. To completely assess motility, the distal tip of the flexible fiberoptic endoscope is withdrawn to just inferior to the aortobronchial compression at which time some bolus trials of various liquid and food consistencies may be conducted to observe the onset of the primary peristaltic wave as well as the clearance effect of the secondary peristaltic wave. Further motility studies may be warranted with high resolution manometry depending upon severity. Once esophageal p.o. trials are completed, final screening for any signs consistent with reflux, candida, etc. can be performed.

#### Prolonged Solid Bolus Fixation Screening

Next, screening is performed to rule out esophageal narrowing with the use of a standardized bolus (e.g. 13 mm placebo pill, or solid food such as 13 mm diced peaches) [44]. In the event a focal prolonged solid bolus fixation greater than 30 seconds with the 13 mm solid bolus is found, then a physiological diagnosis of obstructive esophageal dysphagia is made while referring for a GI physician consultation for definitive medical workup (e.g. EGD with esophageal dilatation and/or biopsy, if indicated, to ascertain and treat the underlying medical etiology of the obstructive esophageal dysphagia).

### P14 = Pharyngeal and Laryngeal Exam with P.O. Trials Augmented with the Clinical Swallow Exam (CSE)

The distal tip of the endoscope is now withdrawn up to the level of the pharynx, and the pharyngeal and laryngeal swallow physiology components of the procedure are performed. This consists of an integration of the Fiberoptic Endoscopic Evaluation of Swallowing (FEES) [45–47]; the Murray Secretion Scale [48]; the Yale Pharyngeal Residue Severity Scale (YPRS) [49,50]; and the Penetration-Aspiration Scale (PAS) [51]. The endoscopist should be aware of the potential impact of the oral and velopharyngeal physiology on pharyngeal swallow function, as well as the influence of comorbidities including but not limited to pharyngoesophageal, esophageal, and gastroesophageal dysphagia. In addition, the effects of comorbid voice disorders, cough disorders, respiratory muscle weakness, dyspneas, and regurgitation (reflux versus backflow versus emesis) upon pharyngeal swallow physiology and collective airway safety of a given patient’s upper aerodigestive tract should be considered. If signs of pharyngeal and/or laryngeal lymphedema are found, then the Revised Patterson Edema Scale is scored [52]. If the patient has a tracheostomy tube, then a subglottic screen photo may be taken looking inferiorly from the posterior glottis to assess airway patency at the level between the glottis and the uppermost portion of the cannula. Furthermore, a lower trachea screening photo may be taken to assess lower airway patency as well as pulmonary toilet by passing the flexible fiberoptic endoscope via the cannula to the distal portion of the trach tube. It is important that clinicians use solid clinical reasoning when introducing p.o. trials during the ADS procedure. For example, if a patient has been strictly nothing by mouth and receiving all intake non-orally (e.g. PEG tube, PEJ tube, etc.), then after an initial visualization of the stomach and the esophagus some p.o. trials may be first initiated with examination at the level of the pharynx. This is to first rule out airway threats occurring immediately before, during, or immediately after the pharyngeal swallow reflex. Once this has been determined, then the clinician may re-introduce the endoscope into the esophagus to complete the esophageal component of the ADS procedure.

### P15 = Re-examination of Esophageal Physiology

Although the gastric screen and the initial esophageal swallow components have been completed prior to initiation of the full pharyngeal-laryngeal exam, it is important to re-examine the stomach and the esophagus as the study progresses in order to differentiate which findings are primarily a result of abnormal pharyngeal and/or laryngeal physiology versus which findings are a primary versus a comorbid synergistic effect from abnormal esophageal and/or gastric findings.

### P16 = Final Velopharyngeal Exam

Prior to removal of the endoscope from the nasal cavity, the velopharynx and nasal cavity are re-examined. As the endoscope is withdrawn, the clinician notes any signs of the color contrast material being expressed on the tissues within or above the velopharyngeal port, adenoidal pad, Eustachian tube orifices, nasal turbinates, etc. which may be additional evidence of velopharyngeal function being grossly impaired.

### P17 = Final Oral Exam

Now a post-ADS oral exam is conducted and documented with a photo(s) for later data analysis of any oral swallow physiology dysfunction which may have occurred during the procedure. Additional visual clinical observations of oral stasis viewed by cues for the patient to open his/her mouth at intervals during the ADS procedure are also noted.

### P18 = Post-ADS Physical Exam

Vital signs are taken again and documented after the procedure is completed. Any adverse effects of the ADS procedure (e.g. mild epistaxis) are reported along with any first aid that was administered. In the event that the patient tolerated the procedure well without any adverse effects, then a statement to this effect is documented into the ADS software for the clinical report.

### P19 = Augmentative/Supplemental Procedures if Applicable

A re-introduction of the endoscope may be performed if needed for any augmentative/supplemental procedures (NMES mapping, sEMG mapping, therapeutic swallow maneuver trials after brief practice without scope in situ, myofascial release trials with follow-up imaging, FEEST, manometry, etc.).

### P20 = Initiation of Data Analysis

As the audiovisual data is reviewed, specific photos as well as brief audiovisual samples of the salient findings are identified and saved to be posted into the ADS clinical report via the ADS software. When a given still photo or brief video file is posted into the storyboard portion of the ADS clinical report, the clinician has the opportunity to write a brief statement of what is salient in the relevant section of the storyboard portion of the report.

### P21 = Completion of Infection Control

At this time, post-procedure infection control protocols are now executed in accordance with government regulations, professional organization guidance, manufacturers guidelines, and local facility protocols.

### P22 = Documentation

Documentation of the completed ADS procedure is accomplished with the use of the ADS software. The ADS software generates a report including a set of digital worksheets used to enhance clinical interpretation of findings from each portion of the aerodigestive tract. The next component of the ADS report is a storyboard which is used to highlight any physiological abnormalities found with accompanying photos and videos to illustrate the interpretation(s) made. Furthermore, each section of the storyboard is used to present the results of any stimulability testing using direct or indirect interventions trialed during the ADS procedure. Lastly, the ADS report summarizes two sets of recommendations. The first set of recommendations are for the safest food, liquid, and oral medication consistencies based upon the ADS findings, as well as recommended methods of safe administration of oral intake (compensatory strategies, swallow maneuvers, positioning of the head, neck, and upper torso, etc). The second set of recommendations are the recommendations for specific evidence-based therapeutic intervention(s), modalities, and manual therapies, for which the patient appeared most stimulable during the procedure. In addition, any recommended specialist consults (e.g. GI, ENT, pulmonology, radiology, physical therapy, etc) are stated along with supporting rationale.

A video covering steps P5 through P17 of the ADS procedure has been added as supplemental material (or available under
https://drive.google.com/drive/folders/1-JFzNGcfcBiOhA-1p4vjKR64g5dASb5R).

### Patient and Public Involvement (PPI) Statement

While patients and public were not involved in the design of the procedure, it was developed with the benefit of the patients and the public in mind to expedite the amount of time from identification of symptoms and signs to diagnosis and appropriate evidence-based treatment recommendations which can represent a gap in care particularly in rural healthcare settings.

## Results

Patient records collected between 2010 and 2021 were assessed for ADS procedure completion. 18,464 ADS procedures were ordered by the referring physicians between 1 January 2011 and 31 December 2021. Of these, 17,881 ADS procedures were completed, while 487 procedures 2.72%) were deemed as aborted procedures which meant the procedure could not be performed as those patients presented as being outside of the vital sign parameters utilized for ensuring patient safety and/or presented with acute signs or symptoms of medical instability or distress. In addition, another 96 (0.53%) procedures were aborted due to patient refusal to permit transnasal passage of the endoscope. For patients unable to complete the procedure, a referral was made back to the ordering physician for further medical evaluation.

Patients were referred for the ADS procedure from physicians, physician assistants, nurse practitioners, nurses, and therapists practicing mostly in rural hospitals, outpatient clinics, and nursing homes across the States of Alabama and Mississippi. Reasons for referrals from physicians for the ADS are summarized Table 1:

**Table 1:** Reasons for ADS referral(s) by chief complaint or clinical rationale

| **Reasons for ADS referral(s)** |  |
| --- | --- |
| **Swallow** | • Difficulty swallowing<br>• Choking/Coughing /Strangling when eating and/or drinking<br>• Globus<br>• Sensation of solid food sticking in throat or in chest<br>• Pain with swallowing<br>• Nasal regurgitation of liquids, foods, and/or medication<br>• Loss of consciousness when swallowing<br>• Requirement of Heimlich Maneuver when eating/drinking |
| **Voice** | • Sudden onset of voice changes especially if persistent<br>• Gradual onset of voice changes<br>• Unfavorable response to swallow therapy<br>• Unfavorable response to voice therapy<br>• Unfavorable response to respiratory therapy especially with failed ventilator weaning attempts and/or failed endotracheal tube extubation attempts<br>• Suspicion for reduced laryngeal and/or tracheal airway patency |
| **Resonance** | • Onset of hypernasality<br>• Onset of hyponasality<br>• Cul de sac resonance<br>• % nasal emissions |
| **Cough** | • Persistent cough<br>• Weakened cough<br>• Cough of unknown origin<br>• Stridor |
| **Respiration** | • Difficulty breathing during or after meals<br>• Difficulty breathing in presence of negative chest X-Ray<br>• Intolerance of one-way tracheostomy speaking valves with tracheostomy patients<br>• Assess readiness for extubation of tracheostomy tubes<br>• Awakening from sleep coughing/ choking |
| **Regurgitation** | • Late-prandial or postprandial vomiting<br>• Suspicion of reflux<br>• H/o of persistent and/or worsening dysphagia |
| **GI** | • Placement of transnasally passed gastric decompression tubes<br>• Placement or placement check of transnasally or transorally placed feeding tubes |
| **Other** | • Fever of unknown origin<br>• Sepsis of unknown origin<br>• Biofeedback therapy session<br>• Follow up to assess progress with pharmaceutical, surgical and/or therapy interventions |

All patients who underwent the ADS procedure were determined to be medically stable by their physicians and were within the ADS vital sign parameters reported in Table 2.

**Table 2:** Recommended vital sign parameters that should be assessed before and after each ADS procedure. Patients outside of these parameters should only be considered under further physician consultation.

| Vital Sign Parameters |  |  |
| --- | --- | --- |
|  | Minimum value | Maximum value |
| Systolic blood pressure | 86 mm Hg | 180 mm Hg |
| Diastolic blood pressure | 34 mm Hg | 120 mm Hg |
| Pulse (bpm) | 40 beats per minute | 120 beats per minute |
| Respiratory rate | 10 breaths per minute | 40 breaths per minute |

### Adverse effects

For the 17,881 ADS procedures completed, no emergency medical personnel were required to manage the occasional adverse effect of mild epistaxis in 109 patients (0.60%). There were no instances of mild vasovagal response (fainting), life threatening laryngospasm, or anaphylaxis. The risk of anaphylaxis was mitigated as no topical anesthesia was used during endoscopy. Furthermore, patient complaints of discomfort (e.g. pressure in the nose, fear of gagging, and actual gagging) were rare.

## Discussion

Here we have described a novel and demonstrably safe endoscopic procedure for the differential diagnosis of primary and secondary physiological deficits of the entire aerodigestive tract from the nasal and oral cavities to the stomach. The ADS procedure aims to optimize patients for healing and the rehabilitation process through identification and management of airway threats which at times can also result in impaired ability to maintain nutrition and hydration. The ADS procedure is unique in that it includes both an esophageal exam and a gastric screen component to identify signs consistent with abnormal gastroesophageal function which, depending upon the underlying etiology, may present a primary or comorbid airway threat.

ADS empowers the diagnostician to perform a comprehensive assessment of all physiological functions of the aerodigestive tract including but not limited to swallow impairments. The robust diagnostic yield of ADS is usually obtainable regardless of a given patient’s level of alertness, cognitive communication status, or seating and positioning limitations. ADS is cost-effective as the initial instrumental assessment of aerodigestive physiological functions especially in rural skilled nursing facilities, outpatient clinics, physician offices, and community hospitals where fluoroscopy, endoscopy, stroboscopy, high resolution manometry, and other advanced instrumentation are often not easily available. The ADS procedure with use of the ADS software typically requires one hour of a diagnostician’s time including set-up, chart review, actual endoscopy time, data analysis, and writing of the report.

Today, videofluoroscopy with the traditional Modified Barium Swallow (MBS) (Logeman, 1983; Logeman, J., 1986) and utilizing the standardized MBS Imp (Martin-Harris, Bonnie, 2015;) are considered the “gold standard” for evaluating swallow physiology. However, the MBS is limited in its current form for screening for GI dysfunction, which at times may be the primary or significant comorbid issue presenting the greatest threat to the airway. Another limitation is that the MBS is unable to provide clearance for the initiation of true vocal fold exercises which may be warranted when impaired glottic closure is the primary reason for aspiration of food, liquid, and oral medications into the trachea. As the MBS is not well suited for ruling out a primary or comorbid GI dysfunction, clinicians using fluoroscopy alone are at risk for misinterpreting the difference between reflux, backflow, and emesis. One other limitation of the MBS is that it is not well suited for identifying comorbid impaired physiologies of the upper aerodigestive tract which may exacerbate a given patient’s dysphagia and/or pose additional threats to airway safety (cough disorders, breathing disorders, reflux disorders, etc.).

Another standard for the assessment of swallow physiology is the Fiberoptic Endoscopic Evaluation of Swallowing (FEES) [45,46]. FEES is well equipped as a follow-up procedure for further evaluation of pharyngeal swallow physiology and/or re-evaluation of pharyngeal swallow physiology after an MBS has been performed. In addition, in healthcare settings where fluoroscopy is unavailable, the FEES procedure administered in conjunction with a clinical swallow exam is well suited for evaluating both oral and pharyngeal swallow physiology as well as their inter-relationship. However, FEES alone, even when administered with a clinical swallow evaluation, is inadequate for the detection of comorbid esophageal dysphagia and gastroesophageal dysphagia. Without a stroboscopic component, it is also ill-equipped for clearing a patient for true vocal fold (TVF) adduction exercises when impaired glottic closure is the primary or comorbid cause of aspiration for a given patient. Although ADS is not intended to replace the MBS, the MBS Imp, or FEES, it has been our experience the performance of the ADS procedure with its software containing guidance for interpretation of the differential diagnostic algorithm has proved successful in identifying reasons why various patient types did not progress with the diagnostic yield of the MBS Imp alone, FEES alone, or even cases when both the MBS Imp and FEES were utilized in conjunction.

Regardless of the chief complaint pertaining to the aerodigestive tract, the holistic approach provided by the ADS procedure for a complete exam of aerodigestive physiology can be utilized to minimize the risk of missing clinically significant comorbidities. The rationale here is similar to that employed in comprehensive differential diagnostic exams utilized in clinical vision science and clinical hearing science. For if each piece of a complex biomechanical system is not carefully examined, then there may be risks for less favorable clinical outcomes especially when all comorbid issues in a given system are not identified early in the diagnostic process. Clear, timely delineation of primary and comorbid physiological deficits of the upper aerodigestive tract is desirable when considering the selection of the most cost-effective holistic treatment(s) particularly given the existing evidence pertaining to cross-over treatment effects [53–58]. Such an approach to identification and management of aerodigestive issues may be considered a superior philosophy of comprehensive airway management across populations. In terms of airway management, there is a sense of urgency for the need of timely identification of comorbid aerodigestive disorders. This is exacerbated in the United States of America by progressively diminished reimbursement with Medicare-dependent as well as privately insured populations, as the number of elderly patients continues to rise each year. Another area of increased utilization in the face of reduced reimbursement is occurring with the increased incidence of late-stage head and neck cancers in the United States of America [59].

The 17,881 patients reported here tolerated this procedure involving unanesthetized transnasal passage of the flexible fiberoptic endoscope to just inside of the stomach without significant complications and minimal discomfort. Patient reports of discomfort included feeling pressure in the nose, elicitation of a cough and/or sneeze, and occasional elicitation of a gag reflex. However, when patients were instructed to concentrate on breathing normally through the nose, they reported a reduction in the sensation of nasal pressure. In addition, for patients who gagged, once cued to breathe in and out of the nose, they reported increased tolerance without gagging. The overall tolerance of the transnasal passage for the ADS procedure appears to be commensurate with other studies examining patient tolerance and comfort levels. Some studies found no statistically significant difference in comfort levels between subjects who received a local anesthesia and subjects who received a placebo for transnasal laryngeal examination procedures such as the FEES procedure [60,61]. Other studies have found transnasal pharyngoesophageal procedures including transnasal esophagoscopy exams and pharyngoesophageal high resolution manometry to be feasible, safe, and well-tolerated by patients [62,63].

The ADS algorithmic protocol has been shown to be comprehensive as it examines the key physiological functions of the entire upper aerodigestive tract most relevant to a given patient’s chief complaint. The protocol is also flexible as it allows the diagnostician to consider the effect(s) of comorbid aerodigestive deficits upon the primary dysphysiological diagnosis of concern. ADS is distinct from other instrumental assessments of physiological functions of the aerodigestive tract as it empowers the diagnostician to identify primary and comorbid physiological impairments from the mouth and nose to the stomach; consider their potential adverse synergistic effects; and develop holistic differential diagnostic statements as well as comprehensive treatment recommendations.

The philosophy of comprehensive airway management at the core of the ADS procedure can be described as follows: 1) to definitively rule out airway threats from above and from below the larynx; 2) to delineate the nature, extent, and severity of disordered physiology of the upper aerodigestive tract; and 3) to assess stimulability for potential evidence-based direct and indirect interventions. For the purposes of this paper, stimulability refers to signs consistent with a favorable prognosis for a specific therapeutic intervention. Indirect interventions include but are not limited to food, liquid, and medication consistency modifications, compensatory strategies (e.g. limited bolus size, cyclic ingestion, slow rate, multiple swallows, etc.), posture modifications (e.g. chin tuck, head rotation, etc.), and maneuvers (e.g. effortful swallow, supraglottic swallow, etc.). Direct interventions include but are not limited to evidence-based therapy interventions (e.g. Lee Silverman Voice Therapy, PhoRTE therapy, McNeil Dysphagia Therapy Program, combined respiratory muscle training, etc.). In addition to assessing stimulability for specific therapeutic interventions during the rehabilitation portion of the ADS procedure, modality trials can also be performed as part of the ADS procedure (e.g. neuromuscular electrical stimulation [NMES] mapping, surface electromyography [sEMG]mapping, etc.) as well as manual therapy trials (e.g. myofascial release, etc.). Lastly, part of the ADS algorithm is designed to identify when there is a need for physician consultation to evaluate and rule out a specific medical etiology as well as physician consultation to evaluate for a specific pharmaceutical intervention. In addition, components are built into the algorithm to identify symptoms and signs warranting a specialty physician or surgeon consultation for further evaluation.

The ADS procedure also affords flexibility in terms of reducing healthcare disparities currently existing between urban healthcare and rural healthcare environments as the latter often do not have immediate access to the armamentarium of procedures readily available in urban areas. The ADS procedure has been successfully deployed across the continuum of rural healthcare settings including community hospitals, outpatient clinics, physician offices, skilled nursing facilities, and home health. Lessons we have learned suggest ADS may also be a cost and time effective diagnostic platform for physiologically based aerodigestive disorders in urban healthcare settings.

Implementation of novel procedures can sometimes be delayed by clinical inertia and reluctance to change [64]. It is interesting to note here one of the seminal publications for the development of the modified barium swallow (MBS) by Dr. Jeirlyn Logemann was published in 1967 [65], and yet the MBS is still not readily available for many residing in rural areas in the USA. Perhaps this reflects the difficulty in updating the human fund of knowledge. Given the potential impact on morbidity and mortality from aerodigestive disorders, we urge clinicians to accept the challenge to integrate this procedure into their diagnostic routine. The reason no single procedure for examining all of the cardinal physiological functions of the upper aerodigestive tract has been reported may reflect upon the upper aerodigestive tract not being considered as a separate and distinct body system in the classical sense (e.g. cardiovascular system, pulmonary system, etc.). Regardless, due to its various influences upon the integrity of airway safety, it warrants being considered as an interconnected synergized system for enabling multiple bodily functions while maintaining comprehensive airway regulation.

## Conclusion

Albeit physically intrusive, aerodigestoscopy (ADS) is a safe, medically non-invasive endoscopic differential diagnostic procedure for physiological disorders of the upper aerodigestive tract. The ADS procedure is guided by an algorithm for the examination of the nasal cavity, oral cavity, pharynx, larynx, trachea, and esophagus, as well as a gastric screen. This procedure allows for physiological diagnosis of the upper aerodigestive tract, while identifying comorbid airway threats from both above and below the level of the larynx. ADS enables the medical team to employ comprehensive airway management to examine a holistic array of treatment options for physiological disorders of the upper aerodigestive tract as guided by the chief complaint. In addition, ADS can prevent further exacerbation of aerodigestive disorders, promote rehabilitation, and maximize overall quality of life. ADS also allows for the rapid identification of comorbid airway threats. Additional research is warranted to further investigate the cost-effectiveness, utility, and the diagnostic yield of ADS against the current standard of diagnostic care for the evaluation of aerodigestive physiological deficits.

## Data Availability

All data produced in the present study are available upon reasonable request to the authors

https://drive.google.com/drive/folders/1-JFzNGcfcBiOhA-1p4vjKR64g5dASb5R

## Acknowledgments

The authors thank Ray E.Boyd, BS for his assistance with the production of the supplemental video demonstrating the endoscopy portion of the ADS procedure on a healthy normal subject. In addition, the authors wish Michael Ard, MD, John Matthew Runnels, MD, Samantha Occipinta, MS, CCC-SLP, and Veda Cochran, RN, BSN for helpful comments and critical reading of the manuscript.

## Disclosures and Contributions

RJA is now the sole owner of Southeastern Biocommunication Associates, LLC which owns the intellectual property rights to the ADS procedure and the ADS software. NB serves as independent Chief Scientist for PN Medical as well as a consulting scientist for Southeastern Biocommunication Associates, LLC.. CG, MM, TM, RM, and AB are consulting scientists for Southeastern Biocommunication Associate, LLC. LJ, SA, CA, and AW declare no conflict of interest.

## Supplemental Material

Esophageal Dysphagia: Helpful terms and concepts.

| Disorder | Definition |
| --- | --- |
| Pharyngoesophageal Dysphagia | Disordered swallow physiology in which normal pharyngeal bolus clearance fails to occur due to interruption of bolus passage from the hypopharynx into the cervical esophagus. NOTE: It is important to ensure impaired hypopharyngeal bolus emptying is not secondary to the presence of severe esophageal stasis. |
| Esophageal Dysphagia (Unspecified) | Disordered swallow physiology resulting from an unspecified impaired bolus flow through the esophagus. |
| Obstructive Esophageal Dysphagia | Disordered esophageal swallow physiology which involves a prolonged solid bolus fixation greater than or equal to 30 seconds with a 13 mm placebo tablet/pill and/or 13 mm diced/cubed peaches resulting from narrowing of the esophageal lumen at one or more specific points along its longitudinal axis. |
| Neurogenic Esophageal Dysphagia | Disordered esophageal swallow physiology which involves a deficit of the primary and/or secondary peristaltic waves resulting in delayed esophageal bolus clearance. This may be with or without tertiary esophageal contractions. |
| Pseudo-obstructive Esophageal Dysphagia | Disordered esophageal swallow physiology in which prolonged bolus fixation < 30 seconds in duration is found to be resulting from edema induced narrowing of the lumen typically of 1/3rd or more of the length of the esophagus. |
| Gastroesophageal Dysphagia | Impaired esophageal swallow physiology in which normal esophageal bolus clearance fails to occur due to the interruption of bolus passage at the Lower Esophageal Sphincter (LES). NOTE: It is important to ensure delayed esophageal emptying is not secondary to severe gastric retention/severely impaired gastric emptying. |

Regurgitation Disorders: Helpful terms and concepts:

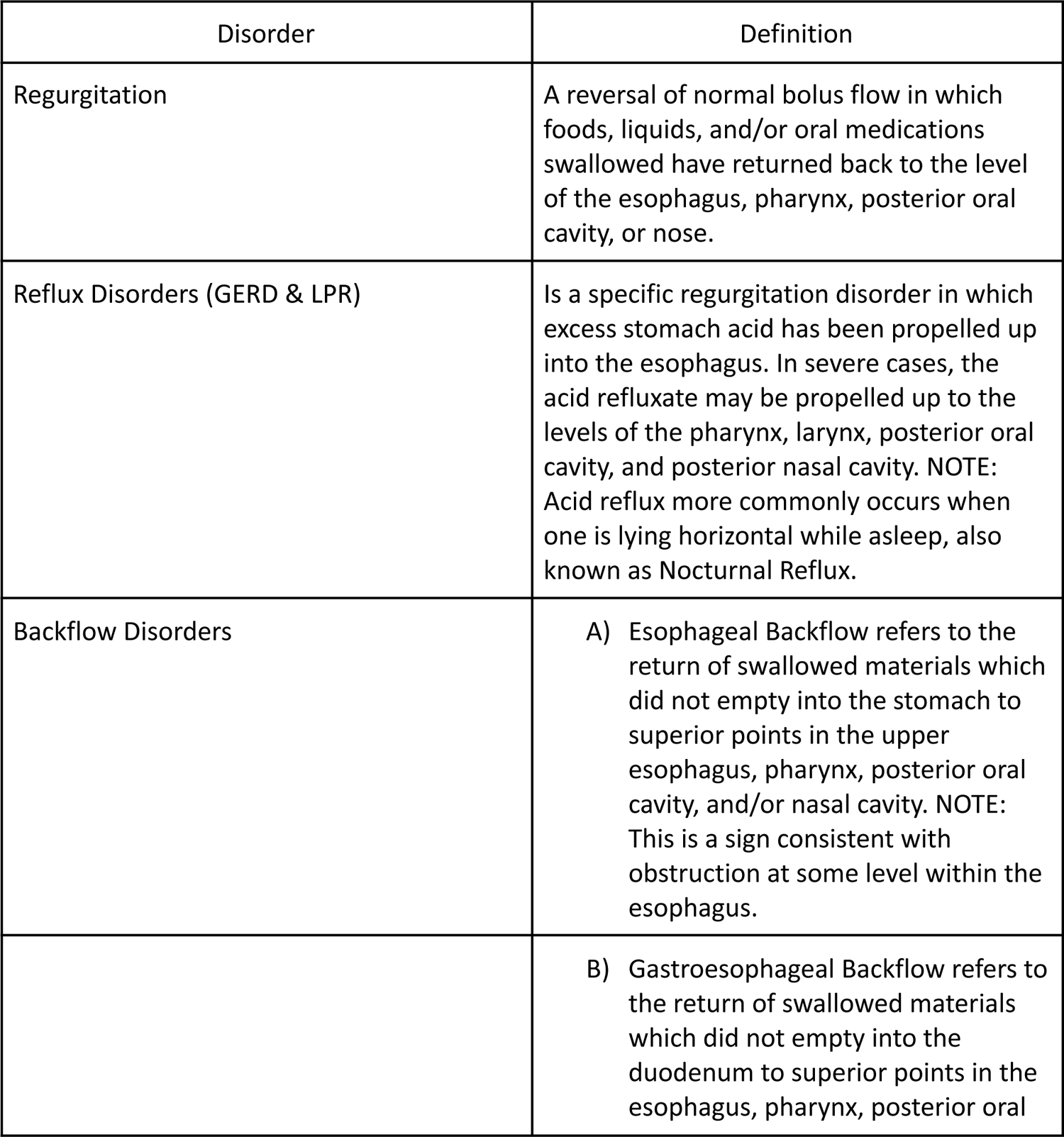
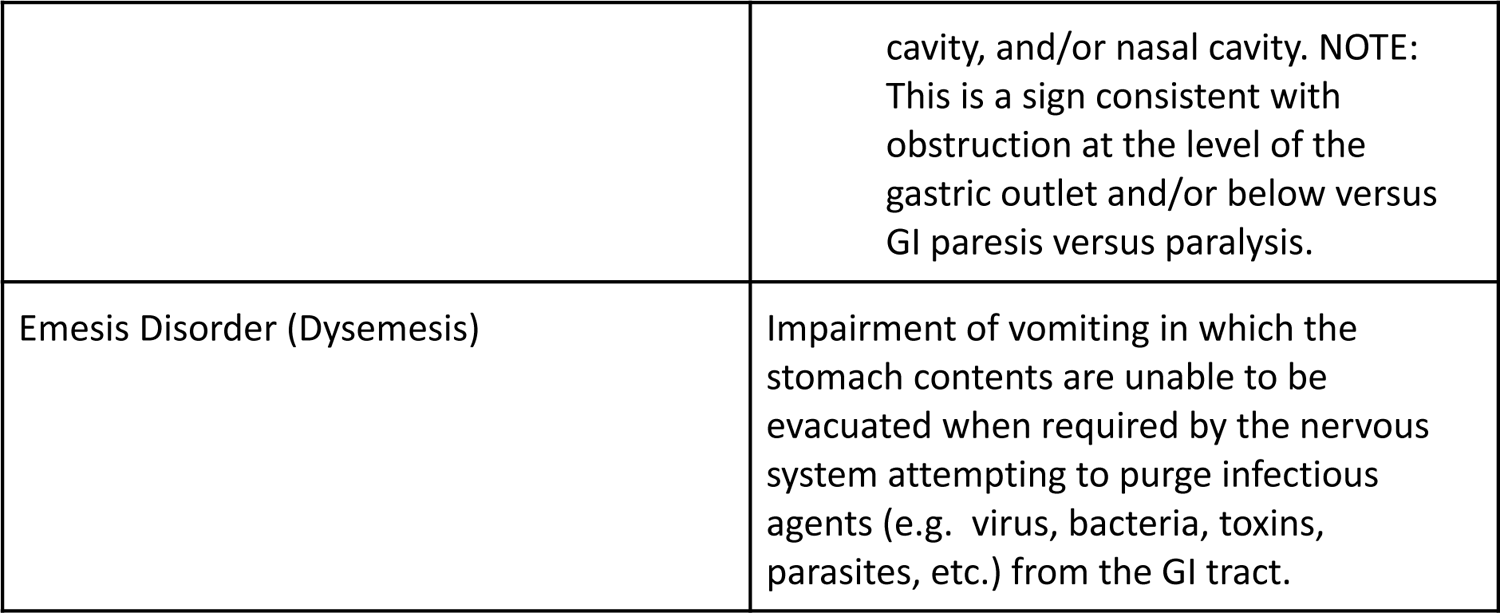

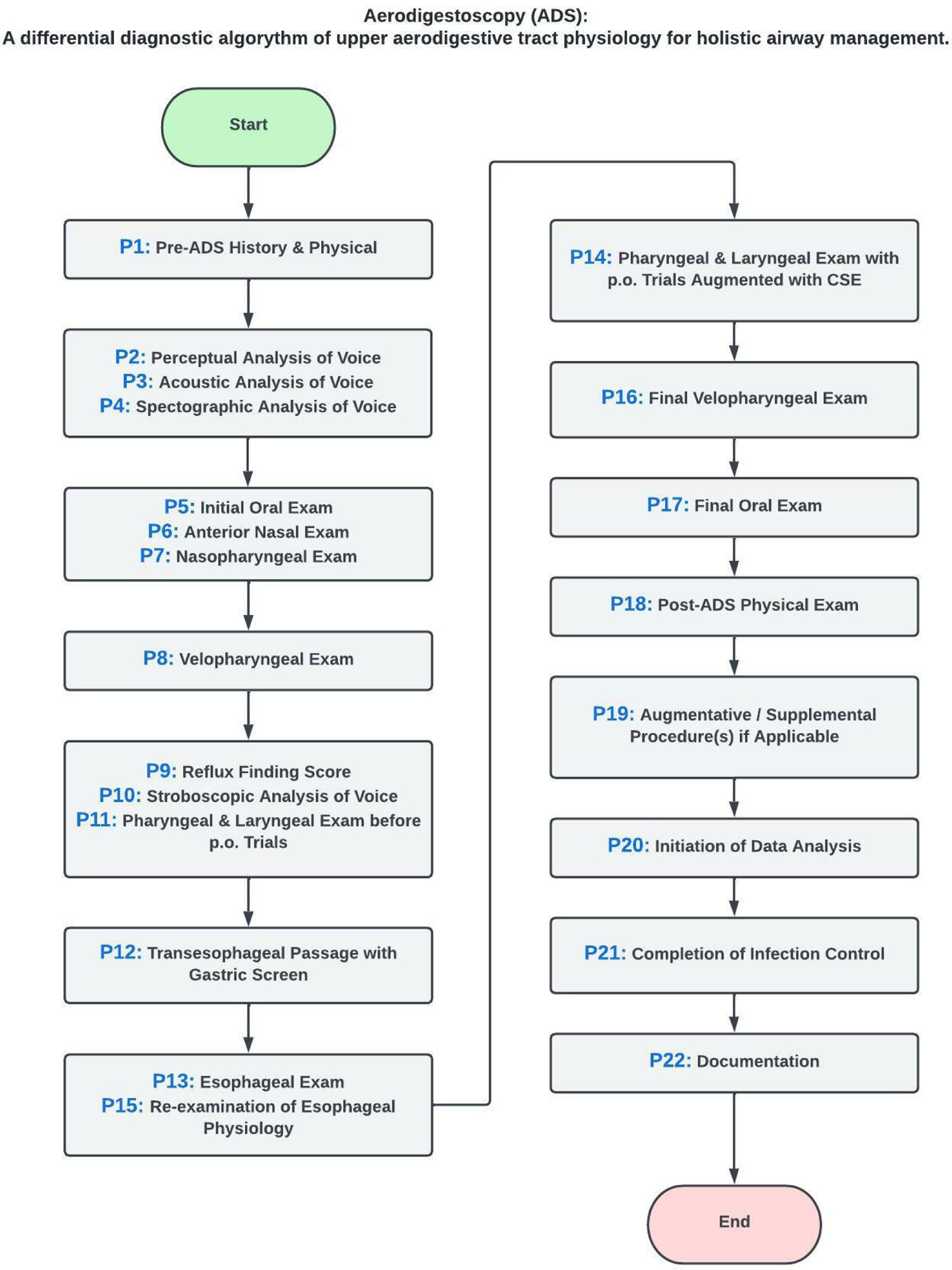

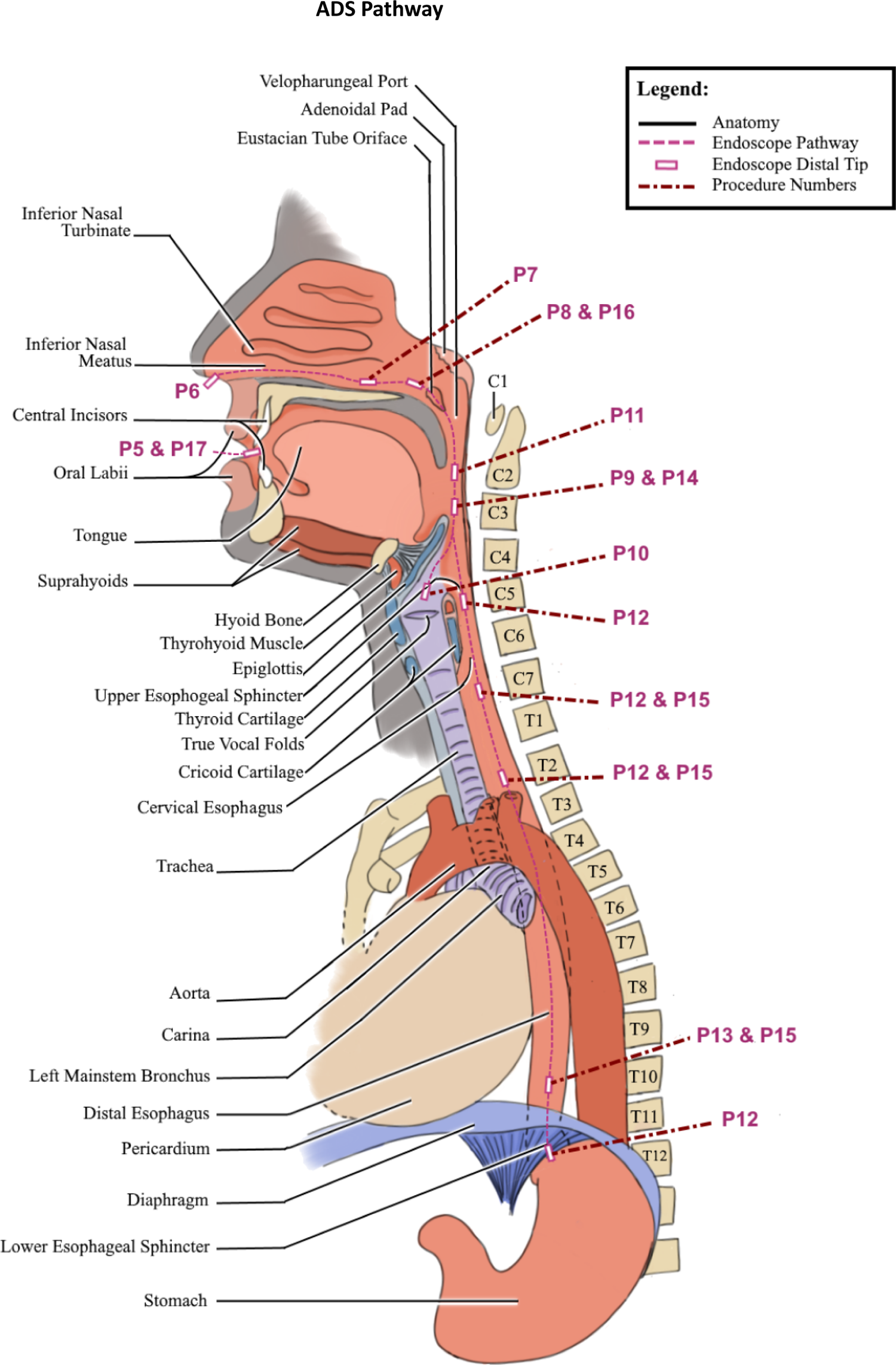

